# Variants in the Niemann-Pick type C gene *NPC1* are not associated with Parkinson’s disease

**DOI:** 10.1101/2020.03.06.20030734

**Authors:** Bouchra Ouled Amar Bencheikh, Konstantin Senkevich, Uladzislau Rudakou, Eric Yu, Kheireddin Mufti, Jennifer A. Ruskey, Farnaz Asayesh, Sandra B. Laurent, Dan Spiegelman, Stanley Fahn, Cheryl Waters, Oury Monchi, Yves Dauvilliers, Alberto J. Espay, Nicolas Dupré, Lior Greenbaum, Sharon Hassin-Baer, Guy A. Rouleau, Roy N. Alcalay, Edward A. Fon, Ziv Gan-Or

## Abstract

Biallelic variants in *NPC1*, a lysosomal gene coding for a transmembrane protein involved in cholesterol trafficking, may cause Niemann-Pick disease type C (NPC). A few cases of *NPC1* mutation carriers have been reported with a Parkinson’s disease (PD) presentation. In addition, pathological studies demonstrated phosphorylated alpha-synuclein and Lewy pathology in brains of NPC patients. Therefore, we aimed to examine whether *NPC1* genetic variants may be associated with PD. Full sequencing of *NPC1* was performed in 2,657 PD patients and 3,647 controls from three cohorts, using targeted sequencing with molecular inversion probes. A total of 9 common variants and 126 rare variants were identified across the three cohorts. To examine association with PD, regression models adjusted for age, sex and origin were performed for common variants, and optimal sequence Kernel association test (SKAT-O) was performed for rare variants. After correction for multiple comparisons, common and rare *NPC1* variants were not associated with PD. Our results do not support a link between heterozygous variants in *NPC1* and PD.

## 1. Introduction

Parkinson’s Disease (PD) is a common neurodegenerative movement disorder, caused by a combination of genetic and environmental factors, with the normal process of aging (Ball et al., 2019; Blauwendraat et al.). The heritability of PD, based on common genetic variance, was calculated in the range of 26-36% (Nalls et al., 2019), yet rare genetic variants also contribute to PD. Numerous genes with common or rare variants associated with PD have a role in the autophagy-lysosome pathway (ALP), including *GBA, LRRK2, SNCA, SMPD1, TMEM175, SCARB2* and others (Senkevich and Gan-Or, 2019).

Variants in several genes that are associated with lysosomal storage disorders have been implicated in PD. Homozygous or compound heterozygous (biallelic) mutations in the glucocerebrosidase (*GBA*) gene result in Gaucher disease, and heterozygous carriers of *GBA* variants are at increased risk for PD (Sidransky et al., 2009). Similarly, biallelic mutations in the lysosomal enzyme gene *SMPD1* may cause Niemann-Pick disease types A and B, and heterozygous variants are associated with risk for PD (Alcalay et al., 2019; Gan-Or et al., 2013). Genetic variants in other genes involved in lysosomal storage disorders, such as *ASAH1, GALC, GLA* and others, may contribute to PD risk as well (Alcalay et al., 2018; Nalls et al., 2019; Robak et al., 2017).

*NPC1* is a lysosomal gene that codes a transmembrane protein involved in cholesterol trafficking (Cruz et al., 2000). Biallelic mutations in *NPC1* may cause Niemann-Pick disease type C, a lysosomal storage disorder with multi-organ involvement (Chiba et al., 2014). Several reported PD cases carrying a mutation in *NPC1* have suggested that *NPC1* mutations could be a risk factor for developing PD (Josephs et al., 2004; Kluenemann et al., 2013; Schneider et al., 2019). The presence of Synuclein in Lewy bodies, the pathological hallmark of PD, have also been documented in post-mortem brain studies of several Niemann-Pick type C patients (Chiba et al., 2014; Saito et al., 2004), further supporting a potential association between *NPC1* mutations and PD.

To study the association between heterozygous variants in the *NPC1* gene and risk for PD, we fully sequenced *NPC1* in a total of 2,657 PD patients and 3,647 controls, and examined the association of both common and rare variants with PD.

## 2. Methods

### 2.1. Population

The study population included 2,657 patients with PD and 3,647 controls from three cohorts, collected at McGill University (Quebec, Canada and Montpellier, France), Columbia University (New York, NY), and Sheba Medical Center (Israel). The McGill cohort includes 1,026 PD patients and 2,588 controls, the Columbia cohort includes 1,026 PD patients and 525 controls, and the Sheba cohort includes 605 PD patients and 534 controls (detailed in Table 1). In all cohorts, to address the age and sex differences, we performed adjustment for age and sex in the statistical analysis. Participants in the McGill cohort were recruited in Québec, Canada (partially through the Quebec Parkinson Network, QPN (Gan-Or et al., 2020)) and in France, and ethnicity were confirmed using principal component analysis. The Columbia cohort has been described previously (Alcalay et al., 2016), and includes patients and controls of mixed ethnicity (European, Ashkenazi [AJ] descent and a minority of Hispanics and blacks). We adjusted for ethnicity when analyzing this cohort. The Sheba cohort was recruited in Israel and includes patients and controls of full AJ origin (all four grandparents are AJ). All three cohorts were sequenced in the same lab (McGill University), following the same protocol. All PD patients were diagnosed by movement disorder specialists according to the UK brain bank criteria (Hughes et al., 1992) or the MDS clinical diagnostic criteria (Postuma et al., 2015)..

**Table 1.** Study population and individuals included in the analysis.

| Sequenced |  |  |  | Analyzed |  |  |  |  |  |
| --- | --- | --- | --- | --- | --- | --- | --- | --- | --- |
|  |  |  |  | Cases |  |  | Controls |  |  |
| Cohort | N<br>(patients) | N<br>(controls) | Depth of<br>Coverage | N | Mean age,<br>(SD) | Male,<br>% | N | Mean age,<br>(SD) | Male,<br>% |
| McGill | 1026 | 2588 | >15x | 973 | 65.5 (10.4) | 62.8 | 2473 | 53.2 (14.22) | 47.2 |
|  |  |  | >50x | 886 | 65.4 (10.3) | 62.2 | 2335 | 53.8 (13.95) | 47.1 |
| Columbia | 1026 | 525 | >15x | 985 | 64.8 (10.9) | 63.9 | 507 | 63.5 (10.2) | 37.1 |
|  |  |  | >50x | 936 | 65.5 (10.9) | 63.9 | 479 | 64.3 (10,0) | 36.7 |
| Sheba | 605 | 534 | >15x | 586 | 63.8 (11.0) | 61.4 | 498 | 33.9 (7.2) | 56.8 |
|  |  |  | >50x | 582 | 63.3 (11.3) | 61.5 | 472 | 33.9 (7.3) | 58.2 |

N, number; SD, standard deviation

### 2.2. Standard Protocol Approvals, Registrations, and Patient Consents

The institutional review board (McGill University Health Center Research Ethics Board - MUHC REB) approved the study protocols (reference number IRB00010120). All local IRBs approved the protocols and informed consent was obtained from all individual participants before entering the study.

### 2.3. Targeted next-generation sequencing by molecular inversion probes

The entire coding sequence of the *NPC1* gene, including exon-intron boundaries (±50bps) and the 5’ and 3’ untranslated regions (UTRs), was targeted using molecular inversion probes (MIPs) as described earlier (O’Roak et al., 2012). All MIPs used to sequence *NPC1* in the present study are included in Supplementary Table 1. Targeted DNA capture and amplification was done as previously described (Ross et al., 2016), and the full protocol is available upon request. The library was sequenced using Illumina HiSeq 2500/4000 platform at the McGill University and Genome Quebec Innovation Centre. Reads were mapped to the human reference genome (hg19) with Burrows-Wheeler Aligner (Li and Durbin, 2009). Genome Analysis Toolkit (GATK, v3.8) was used for post-alignment quality control and variant calling (McKenna et al., 2010), and ANNOVAR was used for annotation (Khan et al., 2019). Data on the frequency of each *NPC1* variant were also extracted from the public database Genome Aggregation Database (GnomAD) (Lek et al., 2016).

### 2.4. Quality control

To only include high-quality variants and samples, we performed quality control (QC) by filtering out variants and samples with reduced quality, using the PLINK software v1.9 (Purcell et al., 2007). Samples with average genotyping rate of less than 87% for minimal coverage 15x and 85% for minimal coverage 50x were excluded. SNPs with genotyping rate lower than 90% were excluded. Common variants that deviated from the Hardy-Weinberg equilibrium set at *p*=0.001 threshold in the control populations were filtered out. Threshold for missingness difference between cases and controls was set at Bonferroni corrected *p*=0.05. To be included in the analysis, the minimum quality score (GQ) of genetic variants was set to 30. Rare variants (minor allele frequency, MAF<0.01) had to have a minimal coverage of >50x to be included, and common variants had to have a minimal coverage of >15x to be included in the analysis. The composition of the three cohorts following these QC steps is described in Table 1.

### 2.5. Statistical Analysis

The association between common *NPC1* variants and PD was examined by using logistic regression models using PLINK v1.9, with the status (patient or control) as a dependent variable, age and sex as covariates in all cohorts, and AJ ancestry as an additional covariate in the NY cohort. To analyze rare variants (MAF < 0.01), an optimized sequence Kernel association test (SKAT-O, R package) (Lee et al., 2012) was performed. SKAT-O was performed for the whole gene, coding and non-coding variants and by comparing synonymous, nonsynonymous, stop frameshift and splicing variants. SKAT-O was also done separately on rare variants with Combined Annotation Dependent Depletion (CADD) score of ≥ 12.37 (Amendola et al., 2015) representing the top 2% of potentially deleterious variants.

## 3. Results

The average coverage of the *NPC1* gene was 880X, with 100% of nucleotides covered at >15X, and 95.7% at >50X in the McGill cohort, 832X, with 100% at >15X and 95,7% at >50X in the Columbia cohort and 1108X, with 100% at >15X and 95.7% at 50X in the Sheba cohort. Supplementary Table 2 details all common variants found in the three cohorts, and their association with risk of PD. None of the common variants was associated with PD in all 3 cohorts after Bonferroni correction (Supplementary Table 2).

**Table 2.** Variants suggested to be involved in PD from previously published studies.

| SNP | nt change | AA change | Distribution in our cohorts | Data from the literature |
| --- | --- | --- | --- | --- |
| rs55680026 | c.A665G | p.Asn222Ser | McGill<br>11 (1.24%) PD / 28 (1.1%) controls | 1 PD patient male 55 age at onset, 65 age at sampling (classical PD) (Zech et al., 2014)<br>CBD-like phenotype AAO- 32, age at sampling 62, main symptoms: depression, later dementia, epilepsy, slight hepatomegaly (Cupidi et al., 2017) |
|  |  |  | Columbia<br>7 (0.75%) PD / 8 (1.67%) controls |  |
|  |  |  | Sheba<br>0 PD / 0 controls |  |
| rs150334966 | c.C3011T | p.Ser1004Leu | McGill<br>3 (0.34%) PD / 7 (0.29%) controls | 2 PD patients with late onset (Age at onset 79 and 87) (Zech et al., 2014) |
|  |  |  | Columbia<br>0 PD / 2 (0.41%) controls |  |
|  |  |  | Sheba<br>0 PD / 0 controls |  |

nt, nucleotide; AA, amino acid

In the McGill cohort, 74 rare variants were included in the analysis, including 26 nonsynonymous variants, 4 splice and 3 loss-of-function variants. In the Columbia cohort, 52 rare variants were included in the analysis, including 16 nonsynonymous variants and 1 splice variant. In the Sheba cohort, 27 rare variants were included in the analysis, including 7 nonsynonymous, 1 splice and 4 loss-of-function variants. In the McGill cohort we found 3 variants that are known as pathogenic for NPC, and 4 variants with conflicting data but likely pathogenic. All these variants except one intronic splice variant were found only in controls. In the Sheba cohort we found 1 likely pathogenic variant for NPC in a control subject. All nonsynonymous, loss-of-function and rare variants affecting splice sites from the three cohorts are detailed in Supplementary Table 3. SKAT-O revealed a nominal association between all rare variants and PD (*p*=0.04) in the McGill cohort, which was mainly driven by synonymous intronic variants. This association was not statistically significant after correction for multiple comparisons. When including only nonsynonymous, splice and loss-of-function variants, or variants with high CADD score, no association was found in any of the cohorts (Supplementary Table 4).

## 4. Discussion

In the current study, we did not find any associations between common and rare variants in *NPC1* and risk of PD, suggesting that *NPC1* does not have a major role in PD in our cohorts. Other lysosomal genes involved in lysosomal storage disorders and in PD, such as *SMPD1, ASAH1, GALC* and potentially *GLA*, are all directly involved in the *GBA* glycosphingolipid metabolism pathway within the lysosome (Senkevich and Gan-Or, 2019). The lack of association with PD of *NPC1* and other genes outside of this pathway (Robak et al., 2017) may suggest that PD may be specifically associated with the *GBA* glycosphingolipid metabolism pathway. *NPC1* is involved in cholesterol trafficking and metabolism; thus far genes from this pathway have not been implicated in PD (Senkevich and Gan-Or, 2019). In our study, all but one of the *NPC1* pathogenic mutations known to cause NPC or potentially pathogenic variants were identified only in controls. Nevertheless, we cannot rule out that very rare disease-causing mutations in *NPC1*, may be associated with PD. Much larger studies will be required to study the role of such mutations in PD.

A recent review (Schneider et al., 2019) summarized cases of patients with neurodegenerative diseases including PD who carried *NPC1* variants. We found in our cohorts a total of 66 carriers of p.Asn222Ser (rs55680026) and p.Ser1004Leu (rs150334966), that have been previously suggested to be associated with PD. Both variants are not very rare and were either equally common or more common in our controls (Table 2). This likely rules out a role for these variants in PD and suggests that their presence in previously reported PD patients may represent chance findings.

Our study has several limitations. In our cohorts, differences between PD patients and controls in sex and age are significant. To address this limitation, we adjusted the regression model with age and sex as covariates. In addition, the cohorts studied here consist of relatively homogeneous populations such as Ashkenazi Jews and French Canadians. Thus, additional studies in other populations are required.

To conclude, our study does not support an important role for common and rare *NPC1* variants in PD. Larger studies will be required to determine whether very rare, specific variants that are pathogenic for Niemann-Pick type C may have a role in PD.

## Data Availability

All data in the manuscript is available upon request

## 5. Acknowledgements

We thank the participants for contributing to the study. This work was financially supported by grants from the Michael J. Fox Foundation, the Canadian Consortium on Neurodegeneration in Aging (CCNA), the Canada First Research Excellence Fund (CFREF), awarded to McGill University for the Healthy Brains for Healthy Lives initiative (HBHL), and Parkinson Canada. The Columbia University cohort is supported by the Parkinson’s Foundation, the National Institutes of Health (K02NS080915, and UL1 TR000040) and the Brookdale Foundation. KS is supported by a post-doctoral fellowship from the Canada First Research Excellence Fund (CFREF), awarded to McGill University for the Healthy Brains for Healthy Lives initiative (HBHL). GAR holds a Canada Research Chair in Genetics of the Nervous System and the Wilder Penfield Chair in Neurosciences. EAF is supported by a Foundation Grant from the Canadian Institutes of Health Research (FDN grant – 154301). ZGO is supported by the Fonds de recherche du Québec - Santé (FRQS) Chercheurs-boursiers award, in collaboration with Parkinson Quebec, and by the Young Investigator Award by Parkinson Canada. OM holds the Canada Research Chair in non-motor symptoms of Parkinson’s disease and the Tourmaline Chair in Parkinson’s disease. The access to part of the participants for this research has been made possible thanks to the Quebec Parkinson’s Network (http://rpq-qpn.ca/en/). We thank Armaghan Alam, Daniel Rochefort, Helene Catoire, Clotilde Degroot and Vessela Zaharieva for their assistance.

## References

Alcalay, R.N., Levy, O.A., Wolf, P., Oliva, P., Zhang, X.K., Waters, C.H., Fahn, S., Kang, U., Liong, C., Ford, B., Mazzoni, P., Kuo, S., Johnson, A., Xiong, L., Rouleau, G.A., Chung, W., Marder, K.S., Gan-Or, Z., 2016. SCARB2 variants and glucocerebrosidase activity in Parkinson’s disease. NPJ Parkinsons Dis 2.

Alcalay, R.N., Mallett, V., Vanderperre, B., Tavassoly, O., Dauvilliers, Y., Wu, R.Y.J., Ruskey, J.A., Leblond, C.S., Ambalavanan, A., Laurent, S.B., Spiegelman, D., Dionne-Laporte, A., Liong, C., Levy, O.A., Fahn, S., Waters, C., Kuo, S.H., Chung, W.K., Ford, B., Marder, K.S., Kang, U.J., Hassin-Baer, S., Greenbaum, L., Trempe, J.F., Wolf, P., Oliva, P., Zhang, X.K., Clark, L.N., Langlois, M., Dion, P.A., Fon, E.A., Dupre, N., Rouleau, G.A., Gan-Or, Z., 2019. SMPD1 mutations, activity, and alpha-synuclein accumulation in Parkinson’s disease. Mov Disord 34(4), 526–535.

Alcalay, R.N., Wolf, P., Levy, O.A., Kang, U.J., Waters, C., Fahn, S., Ford, B., Kuo, S.H., Vanegas, N., Shah, H., Liong, C., Narayan, S., Pauciulo, M.W., Nichols, W.C., Gan-Or, Z., Rouleau, G.A., Chung, W.K., Oliva, P., Keutzer, J., Marder, K., Zhang, X.K., 2018. Alpha galactosidase A activity in Parkinson’s disease. Neurobiol Dis 112, 85–90.

Amendola, L.M., Dorschner, M.O., Robertson, P.D., Salama, J.S., Hart, R., Shirts, B.H., Murray, M.L., Tokita, M.J., Gallego, C.J., Kim, D.S., Bennett, J.T., Crosslin, D.R., Ranchalis, J., Jones, K.L., Rosenthal, E.A., Jarvik, E.R., Itsara, A., Turner, E.H., Herman, D.S., Schleit, J., Burt, A., Jamal, S.M., Abrudan, J.L., Johnson, A.D., Conlin, L.K., Dulik, M.C., Santani, A., Metterville, D.R., Kelly, M., Foreman, A.K., Lee, K., Taylor, K.D., Guo, X., Crooks, K., Kiedrowski, L.A., Raffel, L.J., Gordon, O., Machini, K., Desnick, R.J., Biesecker, L.G., Lubitz, S.A., Mulchandani, S., Cooper, G.M., Joffe, S., Richards, C.S., Yang, Y., Rotter, J.I., Rich, S.S., O’Donnell, C.J., Berg, J.S., Spinner, N.B., Evans, J.P., Fullerton, S.M., Leppig, K.A., Bennett, R.L., Bird, T., Sybert, V.P., Grady, W.M., Tabor, H.K., Kim, J.H., Bamshad, M.J., Wilfond, B., Motulsky, A.G., Scott, C.R., Pritchard, C.C., Walsh, T.D., Burke, W., Raskind, W.H., Byers, P., Hisama, F.M., Rehm, H., Nickerson, D.A., Jarvik, G.P., 2015. Actionable exomic incidental findings in 6503 participants: challenges of variant classification. Genome Res 25(3), 305–315.

Ball, N., Teo, W.-P., Chandra, S., Chapman, J., 2019. Parkinson’s Disease and the Environment. Frontiers in Neurology 10(218).

Blauwendraat, C., Nalls, M.A., Singleton, A.B., The genetic architecture of Parkinson’s disease. The Lancet Neurology.

Chiba, Y., Komori, H., Takei, S., Hasegawa-Ishii, S., Kawamura, N., Adachi, K., Nanba, E., Hosokawa, M., Enokido, Y., Kouchi, Z., Yoshida, F., Shimada, A., 2014. Niemann-Pick disease type C1 predominantly involving the frontotemporal region, with cortical and brainstem Lewy bodies: an autopsy case. Neuropathology 34(1), 49–57.

Cruz, J.C., Sugii, S., Yu, C., Chang, T.Y., 2000. Role of Niemann-Pick type C1 protein in intracellular trafficking of low density lipoprotein-derived cholesterol. J Biol Chem 275(6), 4013–4021.

Cupidi, C., Frangipane, F., Gallo, M., Clodomiro, A., Colao, R., Bernardi, L., Anfossi, M., Conidi, M.E., Vasso, F., Curcio, S.A., Mirabelli, M., Smirne, N., Torchia, G., Muraca, M.G., Puccio, G., Di Lorenzo, R., Zampieri, S., Romanello, M., Dardis, A., Maletta, R.G., Bruni, A.C., 2017. Role of Niemann-Pick Type C Disease Mutations in Dementia. J Alzheimers Dis 55(3), 1249–1259.

Gan-Or, Z., Ozelius, L.J., Bar-Shira, A., Saunders-Pullman, R., Mirelman, A., Kornreich, R., Gana-Weisz, M., Raymond, D., Rozenkrantz, L., Deik, A., Gurevich, T., Gross, S.J., Schreiber-Agus, N., Giladi, N., Bressman, S.B., Orr-Urtreger, A., 2013. The p.L302P mutation in the lysosomal enzyme gene SMPD1 is a risk factor for Parkinson disease. Neurology 80(17), 1606–1610.

Gan-Or, Z., Rao, T., Leveille, E., Degroot, C., Chouinard, S., Cicchetti, F., Dagher, A., Das, S., Desautels, A., Drouin-Ouellet, J., Durcan, T., Gagnon, J.F., Genge, A., Karamchandani, J., Lafontaine, A.L., Sun, S.L.W., Langlois, M., Levesque, M., Melmed, C., Panisset, M., Parent, M., Poline, J.B., Postuma, R.B., Pourcher, E., Rouleau, G.A., Sharp, M., Monchi, O., Dupre, N., Fon, E.A., 2020. The Quebec Parkinson Network: A Researcher-Patient Matching Platform and Multimodal Biorepository. J Parkinsons Dis 10(1), 301–313.

Hughes, A.J., Daniel, S.E., Kilford, L., Lees, A.J., 1992. Accuracy of clinical diagnosis of idiopathic Parkinson’s disease: a clinico-pathological study of 100 cases. J Neurol Neurosurg Psychiatry 55(3), 181–184.

Josephs, K.A., Matsumoto, J.Y., Lindor, N.M., 2004. Heterozygous Niemann-Pick disease type C presenting with tremor. Neurology 63(11), 2189–2190.

Khan, W., Ghani, A., Azmi, M.B., Razzak, S.A., 2019. Beyond sequencing: re-visiting annotations for PJL as a test case. BMC Res Notes 12(1), 467.

Kluenemann, H.H., Nutt, J.G., Davis, M.Y., Bird, T.D., 2013. Parkinsonism syndrome in heterozygotes for Niemann-Pick C1. J Neurol Sci 335(1-2), 219–220.

Lee, S., Emond, M.J., Bamshad, M.J., Barnes, K.C., Rieder, M.J., Nickerson, D.A., Christiani, D.C., Wurfel, M.M., Lin, X., 2012. Optimal unified approach for rare-variant association testing with application to small-sample case-control whole-exome sequencing studies. Am J Hum Genet 91(2), 224–237.

Lek, M., Karczewski, K.J., Minikel, E.V., Samocha, K.E., Banks, E., Fennell, T., O’Donnell-Luria, A.H., Ware, J.S., Hill, A.J., Cummings, B.B., Tukiainen, T., Birnbaum, D.P., Kosmicki, J.A., Duncan, L.E., Estrada, K., Zhao, F., Zou, J., Pierce-Hoffman, E., Berghout, J., Cooper, D.N., Deflaux, N., DePristo, M., Do, R., Flannick, J., Fromer, M., Gauthier, L., Goldstein, J., Gupta, N., Howrigan, D., Kiezun, A., Kurki, M.I., Moonshine, A.L., Natarajan, P., Orozco, L., Peloso, G.M., Poplin, R., Rivas, M.A., Ruano-Rubio, V., Rose, S.A., Ruderfer, D.M., Shakir, K., Stenson, P.D., Stevens, C., Thomas, B.P., Tiao, G., Tusie-Luna, M.T., Weisburd, B., Won, H.H., Yu, D., Altshuler, D.M., Ardissino, D., Boehnke, M., Danesh, J., Donnelly, S., Elosua, R., Florez, J.C., Gabriel, S.B., Getz, G., Glatt, S.J., Hultman, C.M., Kathiresan, S., Laakso, M., McCarroll, S., McCarthy, M.I., McGovern, D., McPherson, R., Neale, B.M., Palotie, A., Purcell, S.M., Saleheen, D., Scharf, J.M., Sklar, P., Sullivan, P.F., Tuomilehto, J., Tsuang, M.T., Watkins, H.C., Wilson, J.G., Daly, M.J., MacArthur, D.G., 2016. Analysis of protein-coding genetic variation in 60,706 humans. Nature 536(7616), 285–291.

Li, H., Durbin, R., 2009. Fast and accurate short read alignment with Burrows-Wheeler transform. Bioinformatics 25(14), 1754–1760.

McKenna, A., Hanna, M., Banks, E., Sivachenko, A., Cibulskis, K., Kernytsky, A., Garimella, K., Altshuler, D., Gabriel, S., Daly, M., DePristo, M.A., 2010. The Genome Analysis Toolkit: a MapReduce framework for analyzing next-generation DNA sequencing data. Genome Res 20(9), 1297–1303.

Nalls, M.A., Blauwendraat, C., Vallerga, C.L., Heilbron, K., Bandres-Ciga, S., Chang, D., Tan, M., Kia, D.A., Noyce, A.J., Xue, A., Bras, J., Young, E., von Coelln, R., Simon-Sanchez, J., Schulte, C., Sharma, M., Krohn, L., Pihlstrom, L., Siitonen, A., Iwaki, H., Leonard, H., Faghri, F., Gibbs, J.R., Hernandez, D.G., Scholz, S.W., Botia, J.A., Martinez, M., Corvol, J.C., Lesage, S., Jankovic, J., Shulman, L.M., Sutherland, M., Tienari, P., Majamaa, K., Toft, M., Andreassen, O.A., Bangale, T., Brice, A., Yang, J., Gan-Or, Z., Gasser, T., Heutink, P., Shulman, J.M., Wood, N.W., Hinds, D.A., Hardy, J.A., Morris, H.R., Gratten, J., Visscher, P.M., Graham, R.R., Singleton, A.B., 2019. Identification of novel risk loci, causal insights, and heritable risk for Parkinson’s disease: a meta-analysis of genome-wide association studies. Lancet Neurol 18(12), 1091–1102.

O’Roak, B.J., Vives, L., Fu, W., Egertson, J.D., Stanaway, I.B., Phelps, I.G., Carvill, G., Kumar, A., Lee, C., Ankenman, K., Munson, J., Hiatt, J.B., Turner, E.H., Levy, R., O’Day, D.R., Krumm, N., Coe, B.P., Martin, B.K., Borenstein, E., Nickerson, D.A., Mefford, H.C., Doherty, D., Akey, J.M., Bernier, R., Eichler, E.E., Shendure, J., 2012. Multiplex targeted sequencing identifies recurrently mutated genes in autism spectrum disorders. Science 338(6114), 1619–1622.

Postuma, R.B., Berg, D., Stern, M., Poewe, W., Olanow, C.W., Oertel, W., Obeso, J., Marek, K., Litvan, I., Lang, A.E., Halliday, G., Goetz, C.G., Gasser, T., Dubois, B., Chan, P., Bloem, B.R., Adler, C.H., Deuschl, G., 2015. MDS clinical diagnostic criteria for Parkinson’s disease. Mov Disord 30(12), 1591–1601.

Purcell, S., Neale, B., Todd-Brown, K., Thomas, L., Ferreira, M.A., Bender, D., Maller, J., Sklar, P., de Bakker, P.I., Daly, M.J., Sham, P.C., 2007. PLINK: a tool set for whole-genome association and population-based linkage analyses. Am J Hum Genet 81(3), 559–575.

Robak, L.A., Jansen, I.E., van Rooij, J., Uitterlinden, A.G., Kraaij, R., Jankovic, J., Heutink, P., Shulman, J.M., 2017. Excessive burden of lysosomal storage disorder gene variants in Parkinson’s disease. Brain 140(12), 3191–3203.

Ross, J.P., Dupre, N., Dauvilliers, Y., Strong, S., Ambalavanan, A., Spiegelman, D., Dionne-Laporte, A., Pourcher, E., Langlois, M., Boivin, M., Leblond, C.S., Dion, P.A., Rouleau, G.A., Gan-Or, Z., 2016. Analysis of DNAJC13 mutations in French-Canadian/French cohort of Parkinson’s disease. Neurobiol Aging 45, 212.e213-212.e217.

Saito, Y., Suzuki, K., Hulette, C.M., Murayama, S., 2004. Aberrant phosphorylation of alpha-synuclein in human Niemann-Pick type C1 disease. J Neuropathol Exp Neurol 63(4), 323–328.

Schneider, S.A., Tahirovic, S., Hardy, J., Strupp, M., Bremova-Ertl, T., 2019. Do heterozygous mutations of Niemann-Pick type C predispose to late-onset neurodegeneration: a review of the literature. J Neurol.

Senkevich, K., Gan-Or, Z., 2019. Autophagy lysosomal pathway dysfunction in Parkinson’s disease; evidence from human genetics. Parkinsonism Relat Disord.

Sidransky, E., Nalls, M.A., Aasly, J.O., Aharon-Peretz, J., Annesi, G., Barbosa, E.R., Bar-Shira, A., Berg, D., Bras, J., Brice, A., Chen, C.M., Clark, L.N., Condroyer, C., De Marco, E.V., Durr, A., Eblan, M.J., Fahn, S., Farrer, M.J., Fung, H.C., Gan-Or, Z., Gasser, T., Gershoni-Baruch, R., Giladi, N., Griffith, A., Gurevich, T., Januario, C., Kropp, P., Lang, A.E., Lee-Chen, G.J., Lesage, S., Marder, K., Mata, I.F., Mirelman, A., Mitsui, J., Mizuta, I., Nicoletti, G., Oliveira, C., Ottman, R., Orr-Urtreger, A., Pereira, L.V., Quattrone, A., Rogaeva, E., Rolfs, A., Rosenbaum, H., Rozenberg, R., Samii, A., Samaddar, T., Schulte, C., Sharma, M., Singleton, A., Spitz, M., Tan, E.K., Tayebi, N., Toda, T., Troiano, A.R., Tsuji, S., Wittstock, M., Wolfsberg, T.G., Wu, Y.R., Zabetian, C.P., Zhao, Y., Ziegler, S.G., 2009. Multicenter analysis of glucocerebrosidase mutations in Parkinson’s disease. N Engl J Med 361(17), 1651–1661.

Zech, M., Nübling, G., Castrop, F., Jochim, A., Schulte, E.C., Mollenhauer, B., Lichtner, P., Peters, A., Gieger, C., Marquardt, T., Vanier, M.T., Latour, P., Klünemann, H., Trenkwalder, C., Diehl-Schmid, J., Perneczky, R., Meitinger, T., Oexle, K., Haslinger, B., Lorenzl, S., Winkelmann, J., 2014. Niemann-Pick C Disease Gene Mutations and Age-Related Neurodegenerative Disorders. PLOS ONE 8(12), e82879.

